# DNA methylation biomarker selected by an ensemble machine learning approach predicts mortality risk in an HIV-positive veteran population

**DOI:** 10.1101/19010272

**Authors:** Chang Shu, Amy C. Justice, Xinyu Zhang, Vincent C. Marconi, Dana B. Hancock, Eric O. Johnson, Ke Xu

## Abstract

**Background:** With the improved life expectancy of people living with HIV (PLWH), identifying vulnerable subpopulations at high risk of mortality is important for clinical care. Evidences showed that DNA methylation (DNAm) is associated with aging and mortality in non-HIV populations. Here, we aimed to establish a panel of DNAm biomarkers that can predict mortality risk among PLWH.

**Methods:** 1,081 HIV-positive participants from the Veterans Aging Cohort Study (VACS) were divided into training (N=460), validation (N=114), and testing (N=507) sets. VACS index was used as a measure of mortality risk among PLWH. Model training and fine-tuning were conducted using the ensemble method in the training and validation sets and prediction performance was assessed in the testing set. The survival analysis comparing the predicted high and low mortality risk groups was conducted. The Gene Ontology enrichment analysis of the predictive CpG sites was performed.

**Results:** We selected a panel of 393 CpGs for the ensemble prediction model. The prediction model showed excellent performance in predicting high mortality risk with an auROC of 0.809 (95%CI: 0.767-0.851) and a balanced accuracy of 0.653 (95%CI: 0.611, 0.693) in the testing set. The predicted high mortality risk group was significantly associated with 10-year mortality (hazard ratio=1.79, p=4E-05) compared with low mortality risk group. These 393 CpGs were located in 280 genes enriched in immune and inflammation responses pathways.

**Conclusions:** We identified a panel of DNAm features associated with mortality risk in PLWH. These DNAm features may serve as predictive biomarkers for mortality risk among PLWH.

## Introduction

Combination antiretroviral therapy has significantly improved the life expectancy of people living with HIV (PLWH), but there is still a gap in life expectancy between PLWH and the general population(1-4). It is important to identify vulnerable groups with a high risk of mortality among PLWH and to deliver early interventions and clinical care for those patients. Previous studies have demonstrated that the Veterans Aging Cohort Study (VACS) index is significantly associated with mortality and is considered a measure of mortality risk among PLWH (5-7). The VACS index is a composite score summing HIV progression measures and general organ injury indicators of the kidneys and liver, which may be able to capture the early stage of elevated risk for mortality.

A large body of evidence has demonstrated that epigenetic modification is influenced by internal and external environmental changes and is associated with the early stages of pathophysiological processes (8-11). DNA methylation (DNAm), one of the most widely studied epigenetic marks, is strongly correlated with aging (12-14), substance use (e.g., cigarette smoking and alcohol consumption) (15-21), and a variety of diseases (8-11, 22, 23). Since DNAm is relatively stable and easy to detect in body fluids obtained through noninvasive procedures, DNAm marks have emerged as robust biomarkers for disease diagnosis (24), disease subtype classification (25, 26) and treatment response monitoring (27, 28).

Since DNAm biomarkers are objectively measured and can reflect pathological processes of disease progression, DNAm can be used to identify individual vulnerability and mortality risk among PLWH. In some cases, DNAm alterations can occur before clinical diagnosis. For example, a longitudinal study of DNAm showed that most DNA methylome changes occurred 80-90 days before clinically detectable glucose elevation (29). As another example, mitochondrial epigenetic changes can indicate early-stage prediabetes (30). Although the clinical diagnosis of myocardial infarction has been well established, DNA methylation in the blood has utility in the diagnosis and monitoring of cardiac pathologies and in the study of normal human cardiac physiology and development (31). These studies support the utility of DNAm features as biomarkers of risk for future onset of disease. Here, we apply this approach to identify individuals with a high risk of mortality in an HIV-positive population.

DNAm plays an important role in HIV infection and disease progression. We previously reported the association of two CpG sites in the promoter region of *NLRC5* with HIV infection (32). *NLRC5* is a major transcriptional activator of the MHC class I gene. DNAm has also been linked to HIV comorbid diseases, such as diabetes and kidney function (33, 34). Furthermore, aging is significantly associated with thousands of CpGs in the epigenome, and the epigenetic clock and DNAm age are becoming widely recognized (12-14). DNAm marks are predictive of mortality in non-HIV populations (35-40). Therefore, we hypothesized that DNAm is associated with mortality risk among PLWH and that DNAm signatures in the blood can serve as biomarkers to predict mortality among HIV-positive individuals.

Machine learning methods have been widely applied to select DNAm features that are informative for the clinical diagnosis and classification of complex diseases (20, 41, 42). Ensemble machine learning methods can aggregate multiple machine learning models (base models) and usually provide better prediction outcomes than single base models (43, 44). An ensemble approach has been shown to perform well in personalized medicine and disease outcomes, such as in cancer and diabetes (45, 46).

In this study, by applying an ensemble machine learning approach, we aimed to identify DNAm features that can serve as biomarkers of mortality risk among HIV-positive individuals. Here, the VACS index was used as a measure of mortality risk in an HIV-positive population (5-7, 47). These predictive DNAm biomarkers can potentially be used for informing future clinical care and providing new insights into the epigenetic mechanism of mortality risk among HIV-positive patients.

## Methods

An overview of our analytical approach is shown in the flowchart in **Figure 1**. Our prediction model was built with training and validation sets profiled on the 450K array and then independently evaluated with the testing set profiled on the EPIC array. Briefly, we first applied an ensemble learning approach to build a machine learning model to predict high or low mortality risk, and we then examined the association of the selected CpG features with mortality by a survival analysis. Then, we conducted a Gene Ontology enrichment analysis for the selected DNAm features. Last, we conducted a meta-analysis of the epigenome-wide association (EWA) on the VACS index of the entire sample.

**Figure 1:**
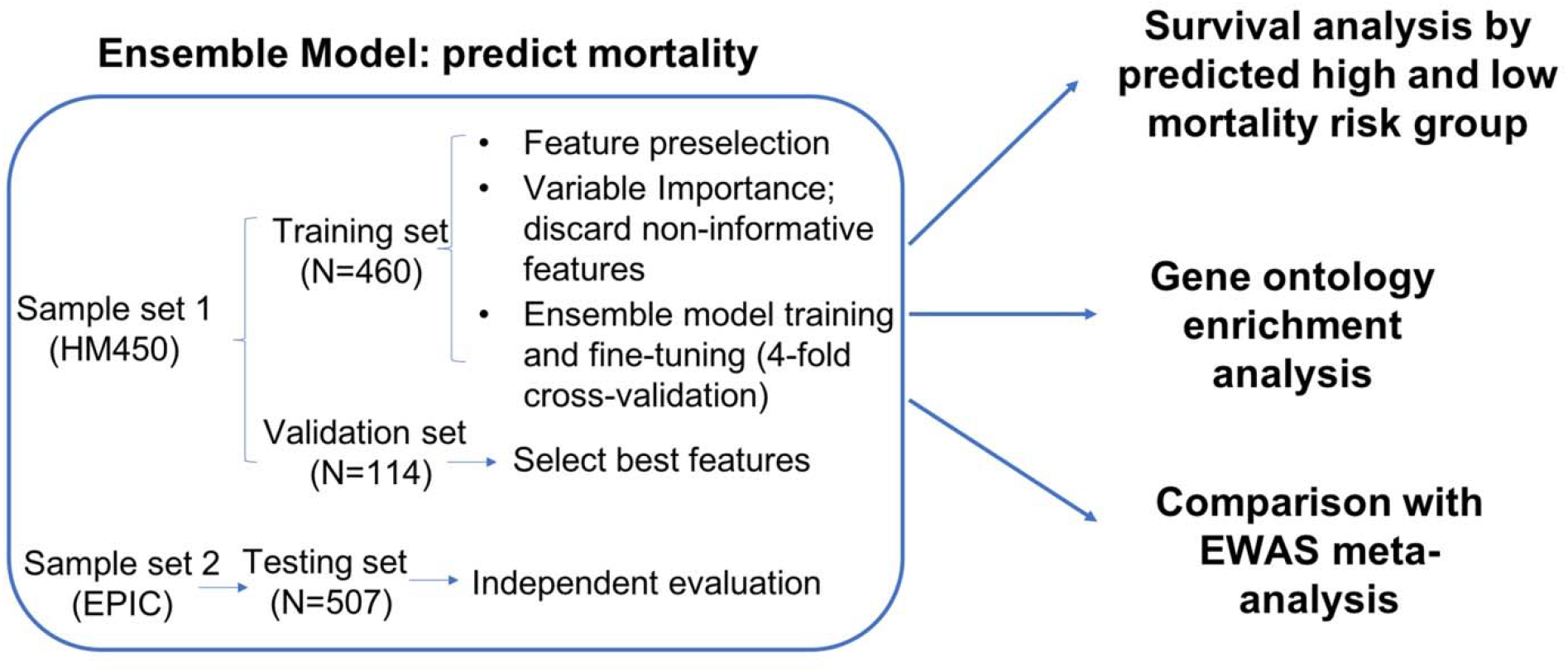
Flowchart of analytical procedures for selecting CpG sites in the peripheral blood methylome, machine learning prediction models to predict high and low mortality risk groups, survival analysis, Gene Ontology enrichment analysis, and epigenome-wide association analysis.

### Study population

All participants in sample sets 1 and 2 (**Figure 1**) were from the VACS that is a prospective cohort study of veterans focusing on the clinical outcomes of HIV infection (5). DNA samples were extracted from the peripheral blood of 1,081 HIV-positive men from the VACS. Participants in sample set 1 were randomly partitioned into a training set (80%, N=460) and a validation set (20%, N=114), and sample set 2 was used as the independent testing set (N=507). Table 1 shows the demographic and clinical information on patient age, sex, race, smoking status, CD4 count, viral load, HIV medication adherence, VACS index, and mortality in the training, validation and testing sets. The training and validation sets included slightly older individuals and more African Americans than the testing set. The VACS index was slightly lower in the testing set than in the training and validation sets. There were no significant differences in sex, smoking, HIV medication adherence, CD4 count, log_10_ HIV-1 viral load or 10-year mortality across the three sample sets.

**Table 1:**
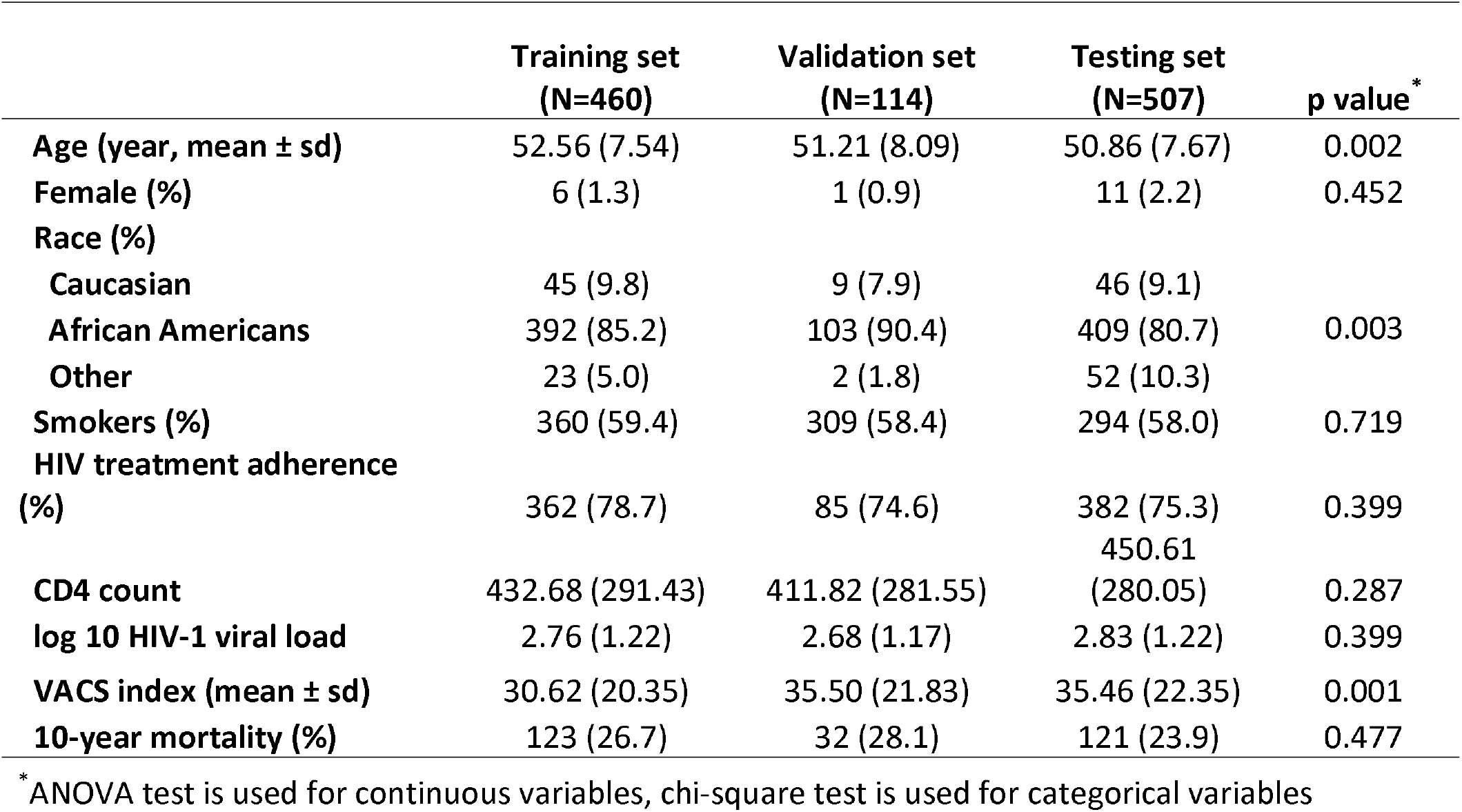
Study sample characteristics.

### Mortality risk

The VACS index is a well-established score for mortality risk among PLWH (5-7). The VACS index is scored by summing preassigned points for age, CD4 count, HIV-1 RNA, hemoglobin, platelets, aspartate and alanine transaminase (AST and ALT), creatine, estimated glomerular filtration rate (eGFR), and viral hepatitis C infection (7). High mortality risk among PLWH was defined as a VACS index score > 40, which was based on a prior observation that the predicted 3-year mortality of 10% for this group was significantly higher than that for the group with VACS index scores ≤ 40(48). Prediction models were developed by machine learning methods to predict high mortality risk (VACS index > 40) and low mortality risk (VACS index ≤ 40) groups among PLWH.

### Genome-wide DNAm profiling and quality control

DNA samples in sample set 1 were profiled by Infinium Human Methylation 450K BeadChip (HM450K, Illumina Inc., CA, USA), and DNA samples in sample set 2 were profiled by the Infinium Human Methylation EPIC BeadChip (Illumina Inc., CA, USA) (**Figure 1**). DNAm for the training and validation sets were evaluated using the same quality control (QC) protocol (49) in the R package *minfi* (50). In detail, CpG sites on sex chromosomes and within 10 base pairs of a single nucleotide polymorphism were removed. The detection p-value threshold was set at 10^-12^ for both sample sets 1 and 2. After QC, 408,583 CpG sites common between the HM450K and EPIC arrays were used for analysis to ensure the same coverage between the two sets. DNA methylation among the common CpG sites was highly correlated between the HM450K and EPIC arrays (r=0.986). Proportions of 6 cell types (CD4+ T cells, CD8+ T cells, natural killer T cells, B cells, monocytes and granulocytes) were estimated for all participants in sample sets 1 and 2 using an established method (51).

### Feature selection of CpG sites in the training set

We first preselected a panel of CpG sites associated with high mortality risk among PLWH based on the EWA of the VACS index score in the training set. CpG sites with p<0.001 were preselected to build the prediction models. A liberal cutoff of p<0.001 was arbitrarily set to ensure a sufficient number of predictive DNAm features to build the prediction models. The variable importance (a score between 0-100) of each preselected CpG site was ranked by elastic-net regularized generalized linear models (GLMNET) by the R package *caret* (52) based on 100 bootstraps, where each bootstrap included 70% of all samples. CpG sites with zero variable importance for 80% of the bootstraps were considered to be nonpredictive features and were removed from further model development. The remaining CpG sites were ranked based on the median importance ranking among 100 bootstraps and were divided into 20 groups. Each CpG group was used to build machine learning models.

### Developing machine learning prediction models for mortality risk among PLWH

1) Model development in the training set: We developed an ensemble method that aggregated the prediction results from four base machine learning models: random forest (RF), GLMNET, support vector machines (SVM) and k-nearest neighbors (k-NN) (53-56) using the model choice of “rf”, “glmnet”, “svmLinear” and “knn” in the R package *caret* (52). These four base models have been commonly used in predicting binary outcomes and have expanded the diversity of algorithms (53-56). Ten-fold cross-validation was used in the model training process to minimize overfitting. These four base models were independently trained to predict mortality risk among PLWH in the training set and then aggregated by the ensemble method using the R package *caretEnsemble* (ver. 2.0.1) (57). The prediction performance of each ensemble model was evaluated by using area under the receiver operating characteristic curve (auROC) and the area under the precision-recall curve (auPRC).

2) Final CpG group selection in the validation set: The CpG group with the highest auROC in the validation set was selected as the final feature group for the ensemble model.

3) Independent evaluation in the testing set: Using the ensemble model and the final feature group, we predicted the high mortality risk group and evaluated prediction performance in the testing set by using auROC and balanced accuracy. Balanced accuracy was defined as the average accuracy obtained on each class, as shown in the following formula (58). Balanced accuracy was used in this study to avoid biased accuracy due to imbalanced samples (58). The 95% confidence interval of balanced accuracy was estimated by 1,000 stratified bootstraps of the testing set.

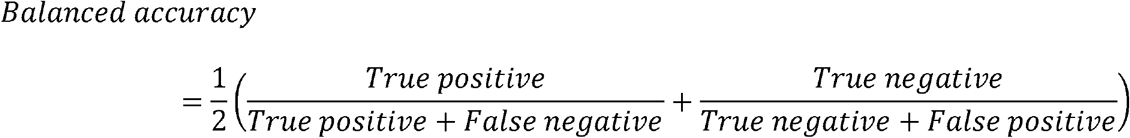

### Survival analysis

By using the final ensemble model, we classified each individual in the entire sample as having as high or low mortality risk. Kaplan-Meier survival curves presented 10-year survival probability by high or low mortality risk group. Survival analysis was conducted using a Cox proportional hazards model on 10-year mortality comparing the high and low mortality risk groups. We used age as time scale *t*, and our model adjusted for sex, race, smoking, self-reported HIV medication adherence, log_10_ of HIV viral load and CD4 count.

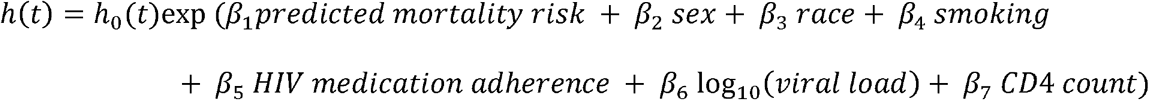

### Biological interpretation of the predictive panel of CpG sites on mortality risk among PLWH

We performed Gene Ontology (GO) enrichment analysis using *missMethyl* to adjust for bias by different numbers of CpG sites per gene (59, 60). Genes that harbor or are located near at least one predictive CpG site were used for GO analysis.

### Epigenome-wide association analysis on mortality risk among PLWH in all samples

Since DNA methylation of two sample sets was measured by two different platforms, we performed EWA on high and low mortality risk groups separately in sample sets 1 and 2, and we then conducted a meta-analysis to detect epigenome-wide signals in the entire sample. In each EWA, we used a two-step linear model approach as previously described (49). The EWA model adjusted for confounding factors, including age, sex, race, smoking, cell type proportions and control principle components. EWA meta-analysis of sample sets 1 and 2 was conducted using METAL (61). The weights of effect size were the inverse of the corresponding standard errors for the meta-analysis (61). CpG sites with Bonferroni corrected p-value < 0.05 were considered statistically significant.

## Results

### Feature selection and ensemble model training for mortality risk among PLWH

High mortality risk among PLWH was defined as VACS index > 40 based on previous literature showing a predicted 3-year mortality of 10% for this group (48). Prediction models were developed by machine learning methods to predict high mortality risk (VACS index > 40) and low mortality risk groups (VACS index ≤ 40) among PLWH.

A panel of 856 CpGs with p<0.001 was preselected based on EWA in the training set. We ranked these candidate predictors by median GLMNET importance ranking among 100 bootstraps using 70% of the training sample. We excluded 178 CpG sites that had a variable importance score of zero among 80% of the bootstraps. A final panel of 678 CpG sites were selected and formed into 20 groups based on importance ranking to determine the best performing CpG group for the ensemble model (**Figure 2**).

**Figure 2:**
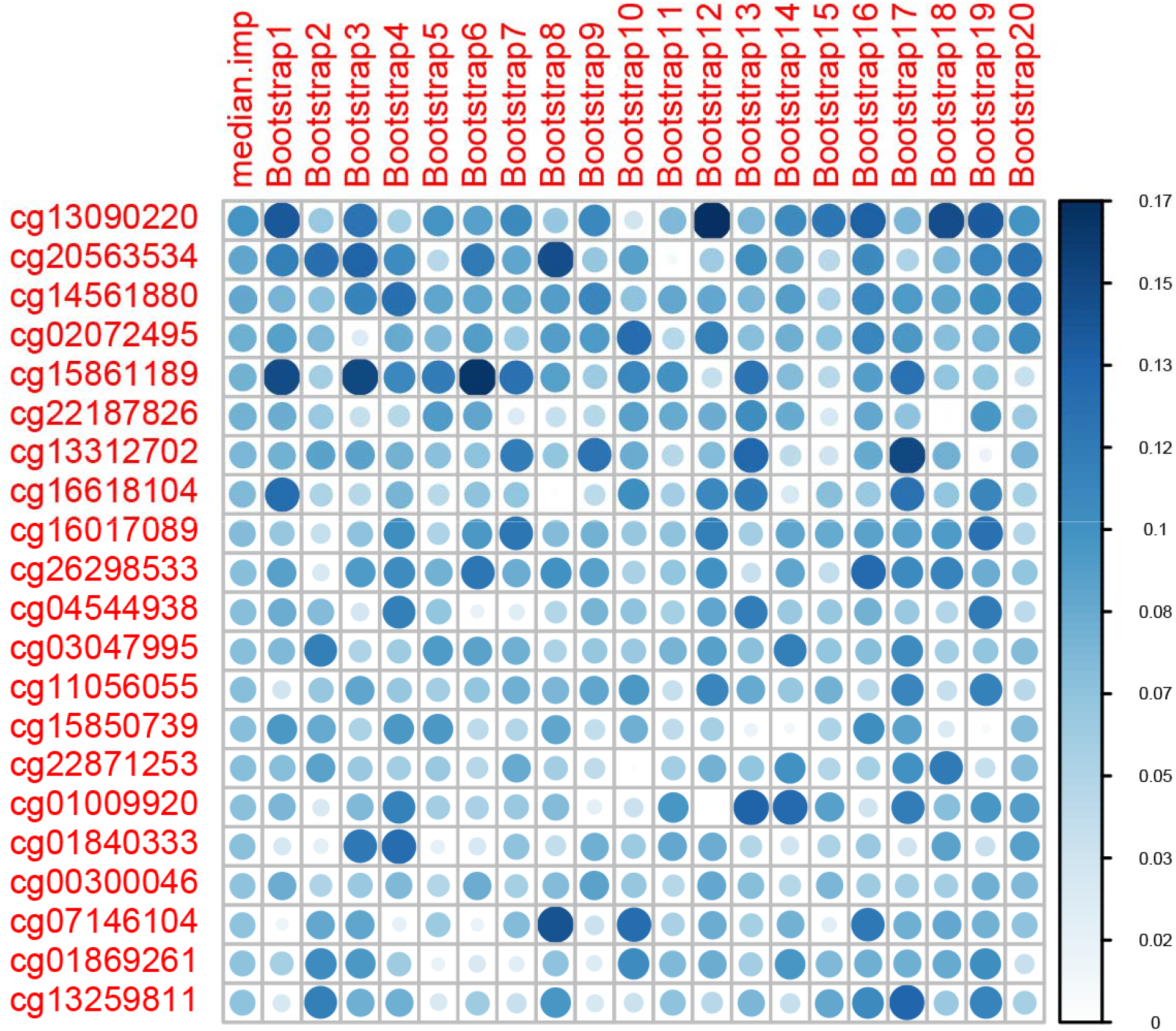
Variable importance ranking of predictive machine learning CpG sites. Variable importance is a score between 0 and 100, as calculated by elastic-net regularized generalized linear models (GLMNET). We obtained variable importance scores from 100 bootstraps. The top 20 ranked CpG sites and 20 bootstraps are shown.

In the training set, we used 4 common machine learning classification models (RF, GLMNET, SVM, and k-NN) as our base models for the ensemble method (53-56) and trained them independently for the 20 groups of CpGs. The performance metrics of GLMNET, RF and SVM were mostly comparable in terms of auROC and auPRC, and they plateaued to 1 with an increasing number of CpGs in the training set (**Figure 3**, **Figure 4**). The performance of k-NN was poorer than the other 3 methods, but its auROC and auPRC remained above 0.9 in the training set. An ensemble model combining the prediction results of all 4 base models was used (**Figure 3**, **Figure 4**).

**Figure 3:**
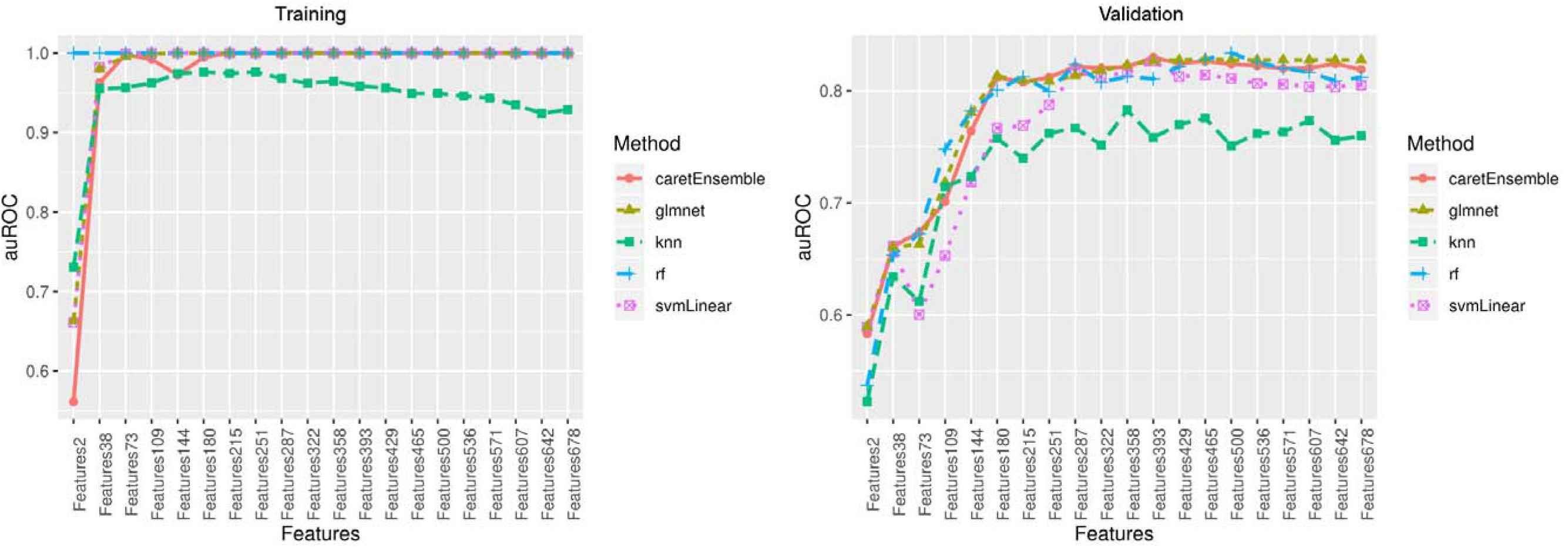
Area under the receiver operating characteristic (auROC) curve in the training and validation sets.

**Figure 4:**
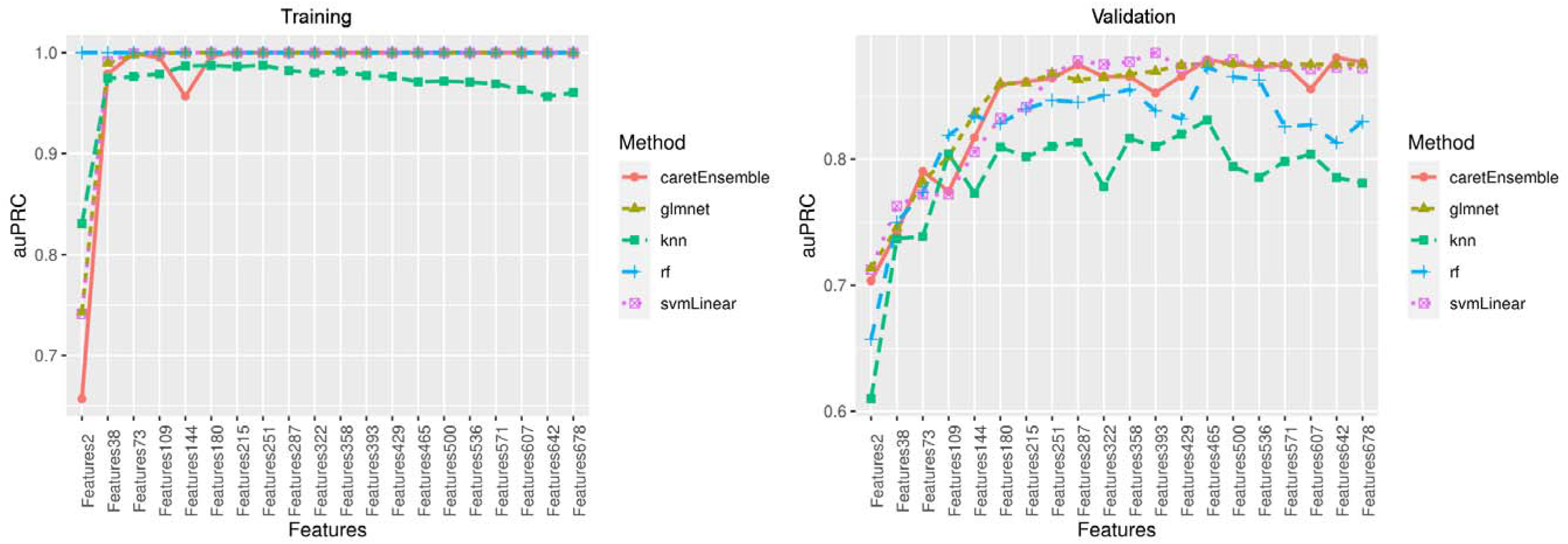
Area under the precision-recall curve (auPRC) in the training and validation sets.

In the validation set, the performances of 4 base models varied. Three models, GLMNET, RF, and SVM, showed good performance with both auROC and auPRC > 0.8, but the performance of k-NN was poor with auROC < 0.8 (**Figure 3**, **4**). The ensemble model showed the best performance at 393 CpG sites with an excellent performance of auROC (0.829). The accuracy of this prediction model was 0.807, and the balanced accuracy that accounted for class imbalances between participants at high and low risk of mortality was 0.782. Thus, the ensemble model with 393 DNAm features was used as the final prediction model (**Table S1**).

In the testing set, the ensemble model with 393 DNAm features showed excellent performance with auROC of 0.809 (95%CI: 0.767-0.851), prediction accuracy of 0.761 and balanced accuracy of 0.653 (95%CI: 0.611, 0.693) (**Figure 5**), suggesting that our ensemble model with a panel of 393 features was able to differentiate between high and low mortality risk in an HIV-positive population.

**Figure 5:**
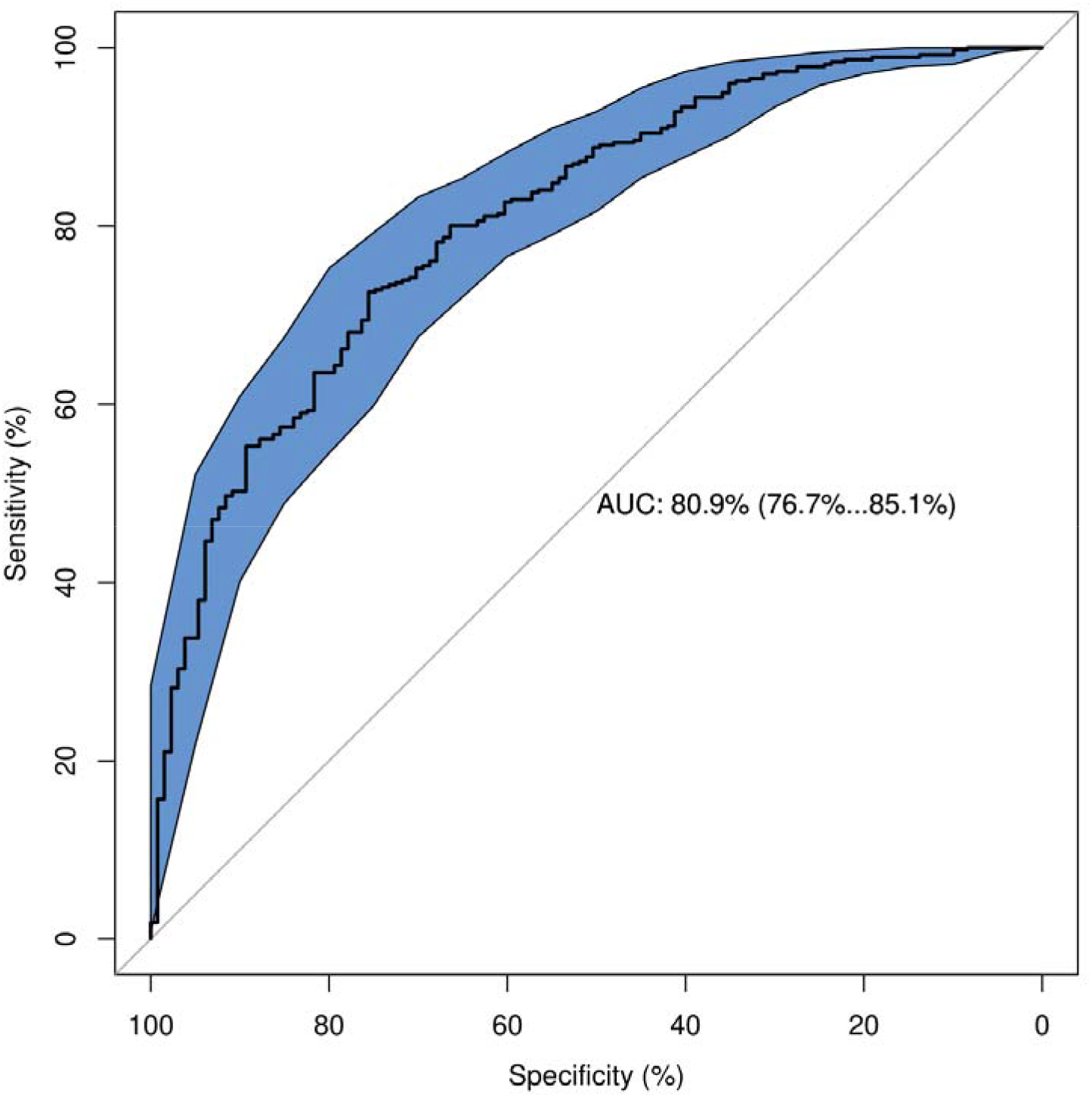
Receiver operating characteristic (auROC) curve in the testing set.

### Survival analysis

By using our ensemble model that predicts individual mortality risk, we found that participants predicted to have a high risk of mortality showed significantly lower 10-year survival probability than participants predicted to have a low risk of mortality (**Figure 6**). Using a Cox proportional-hazards model adjusting for baseline age, sex, race, viral load, CD4 count and antiviral medication adherence, participants who were predicted to have a high risk of mortality remained to have an elevated risk of mortality compared with those predicted to have a low risk of mortality, with a hazard ratio (HR) of 1.79 (95%CI: 1.35-2.37, p=4E-05).

**Figure 6:**
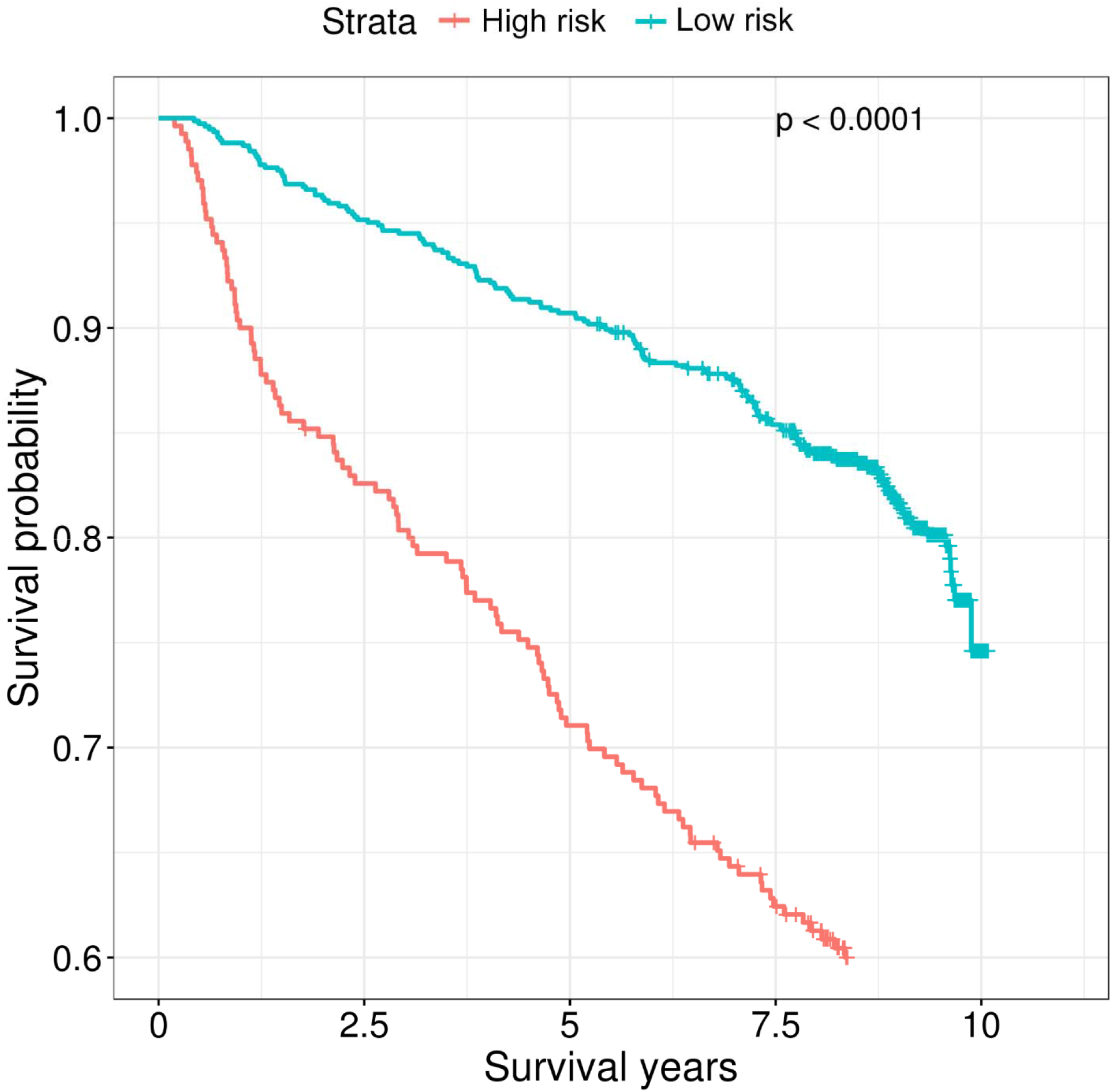
Kaplan-Meier curves of predicted high and low mortality risk groups among people living with HIV.

### Biological interpretations of predictive CpGs by Gene Ontology enrichment analysis

The 393 predictive CpGs were located in or near 280 genes. The top 8 enriched pathways based on these 280 genes included response to virus (p= 4.26E-05), defense response (p=1.29E-04), cytokine receptor binding (p=1.48E-04) and regulation of response to interferon-gamma (p= 4.15E-04) (**Table 2**). Our findings suggested that the selected 393 CpG sites are biologically relevant to HIV pathogenesis and progression.

**Table 2:**
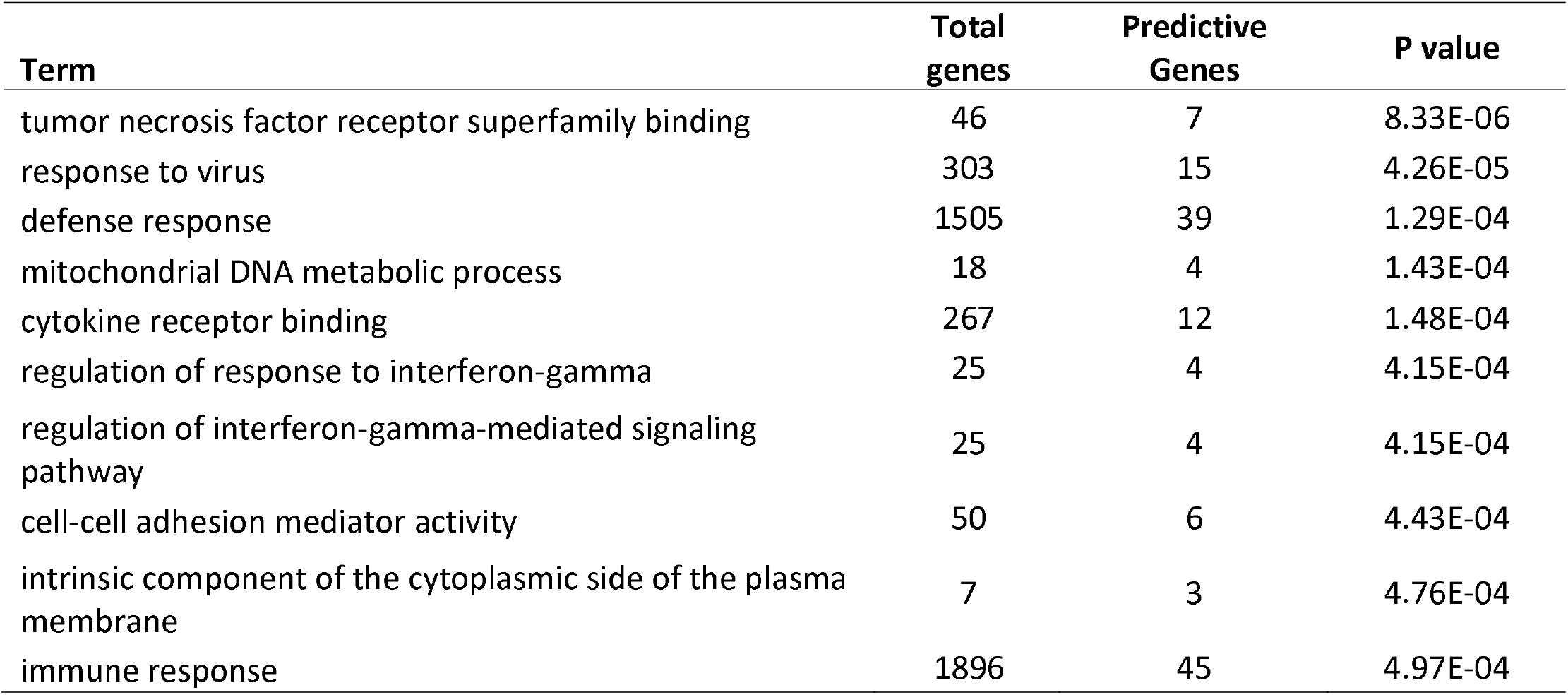
Gene ontology term enrichment analysis of the selected 393 CpG sites that predict mortality risk among HIV-positive population.

| **Term** | **Total genes** | **Predictive Genes** | **P value** |
| --- | --- | --- | --- |
| tumor necrosis factor receptor superfamily binding | 46 | 7 | 8.33 E-06 |
| response to virus | 303 | 15 | 4.26E-05 |
| defense response | 1505 | 39 | 1.29E-04 |
| mitochondrial DNA metabolic process | 18 | 4 | 1.43E-04 |
| cytokine receptor binding | 267 | 12 | 1.48E-04 |
| regulation of response to interferon-gamma | 25 | 4 | 4.15 E-04 |
| regulation of interferon-gamma-mediated signaling pathway | 25 | 4 | 4.15 E-04 |
| cell-cell adhesion mediator activity | 50 | 6 | 4.43 E-04 |
| intrinsic component of the cytoplasmic side of the plasma membrane | 7 | 3 | 4.76E-04 |
| immune response | 1896 | 45 | 4.97E-04 |

### Epigenome-wide association on mortality risk among PLWH

A meta-analysis of EWA of sample sets 1 and 2 identified 208 epigenome-wide significant CpGs after Bonferroni correction (**Figure S2**). These significant CpG sites were located in 112 genes and included genes previously reported as being associated with HIV pathogenesis. For example, cg07839457 in *NLRC5* was previously reported to be associated with HIV infection (32). Interestingly, 30 of 208 CpG sites were also in the panel of machine learning predictive CpGs (**Table 3**). Twenty out of the 30 overlapping CpG sites were negatively associated with mortality risk, while 10 CpG sites were positively associated with mortality risk. Some of the overlapping CpGs were located in viral response genes, such as *IFITM1* and *PARP9*.

**Table 3:** Overlapping CpG sites between machine learning selected CpG sites and epigenome-wide significant CpG sites on mortality risk among people living with HIV.

| probe | Chr | Position | Nearest gene | Variable Importance | Meta Effect (SE) | Meta P | Refgene group | Relation to CpG island |
| --- | --- | --- | --- | --- | --- | --- | --- | --- |
| cg01971407 | 11 | 313624 | *IFITM1* | 9.9 | -0.0399 (0.0039) | 8.05E-25 | TSS1500 | N_Shelf |
| cg22930808 | 3 | 122281881 | *PARP9;DTX3L* | 13.7 | -0.1057 (0.0105) | 7.24E-24 | 5UTR;TSS1500 | N_Shore |
| cg23570810 | 11 | 315102 | *IFITM1* | 13.0 | -0.0638 (0.0065) | 1.15E-22 | Body | N_Shore |
| cg14864167 | 8 | 66751182 | *PDE7A* | 12.0 | -0.0832 (0.0085) | 1.54E-22 | Body | N_Shelf |
| cg01190666 | 20 | 62204908 | *PRIC285* | 19.4 | -0.0343 (0.0035) | 1.79E-22 | 5UTR | N_Shore |
| cg11702942 | 8 | 144102584 | *LY6E;LY6E* | 7.8 | -0.0382 (0.004) | 7.52E-22 | Body;Body | S_Shore |
| cg03607951 | 1 | 79085586 | *IFI44L* | 23.0 | -0.0683 (0.0072) | 3.58E-21 | TSS1500 |  |
| cg03848588 | 9 | 32525008 | *DDX58* | 14.4 | -0.0274 (0.0029) | 6.02E-21 | Body | N_Shore |
| cg04582010 | 11 | 313120 | *IFITM1* | 22.1 | -0.0455 (0.0052) | 2.19E-18 | TSS1500 | S_Shore |
| cg18394552 | 5 | 159428643 |  | 24.4 | 0.0342 (0.0043) | 1.57E-15 |  |  |
| cg03753191 | 13 | 43566902 | *EPSTI1;EPSTI1* | 4.1 | -0.018 (0.0023) | 2.43E-15 | TSS1500 | S_Shore |
| cg17267239 | 1 | 173640200 | *ANKRD45* | 21.8 | -0.0186 (0.0025) | 1.22E-13 | TSS1500 | S_Shore |
| cg12461141 | 11 | 5710654 | *TRIM22* | 37.6 | -0.0274 (0.0037) | 1.37E-13 | TSS1500 |  |
| cg09251764 | 17 | 6659070 | *XAF1;XAF1* | 13.8 | -0.0142 (0.002) | 3.93E-13 | TSS200 |  |
| cg05626226 | 4 | 106515450 | *FLJ20184* | 11.5 | 0.0279 (0.004) | 2.45E-12 | Body |  |
| cg22959742 | 10 | 13913931 | *FRMD4A* | 16.4 | 0.0289 (0.0041) | 3.08E-12 | Body |  |
| cg16936953 | 17 | 57915665 | *TMEM49* | 11.3 | -0.0379 (0.0056) | 1.95E-11 | Body |  |
| cg18181703 | 17 | 76354621 | *SOCS3* | 32.6 | -0.0215 (0.0033) | 1.15E-10 | Body | N_Shore |
| cg07107453 | 1 | 79114976 | *IFI44* | 4.8 | -0.0292 (0.0046) | 2.26E-10 | TSS1500 |  |
| cg25114611 | 6 | 35696870 | *FKBP5;LOC285847* | 12.8 | -0.0125 (0.002) | 4.78E-10 | TSS1500;Body | S_Shore |
| cg01059398 | 3 | 172235808 | *TNFSF10* | 26.7 | -0.0289 (0.0047) | 7.85E-10 | Body |  |
| cg19459791 | 15 | 65363022 |  | 15.1 | 0.0159 (0.0026) | 1.20E-09 |  | S_Shelf |
| cg26282236 | 12 | 1025755 | *RAD52* | 30.2 | 0.0243 (0.0041) | 3.63E-09 | Body |  |
| cg06357748 | 12 | 1025529 | *RAD52* | 10.7 | 0.0277 (0.0047) | 4.12E-09 | Body |  |
| cg04442417 | 20 | 62191507 | *PRIC285;PRIC285* | 26.6 | 0.0201 (0.0035) | 1.22E-08 | Body | Island |
| cg14602222 | 12 | 1025663 | *RAD52* | 13.2 | 0.0241 (0.0042) | 1.42E-08 | Body |  |
| cg26724018 | 11 | 5716255 | *TRIM22* | 11.9 | -0.0171 (0.003) | 1.49E-08 | 5UTR |  |
| cg03084350 | 3 | 38065265 | *PLCD1;PLCD1;PLCD1* | 18.0 | 0.0114 (0.002) | 2.05E-08 | Body | N_Shore |
| cg00569896 | 4 | 204382 |  | 0.2 | 0.0254 (0.0047) | 5.78E-08 |  | N_Shore |
| cg12126344 | 1 | 12207564 |  | 8.4 | -0.011 (0.002) | 7.36E-08 |  |  |

## Discussion

In this study, we presented evidence that DNAm marks in blood were predictive of mortality risk in an HIV-positive population. We identified a panel of 393 CpG sites that were highly predictive for high vs. low risk of mortality among PWLH. We also found that our predicted mortality risk based on the ensemble model was strongly associated with mortality in HIV-positive individuals in the VACS cohort. In addition, the selected 393 DNAm features were located in genes enriched in HIV pathogenesis and progression. Thus, we identified a panel of 393 DNAm biomarkers that may enhance the understanding of the epigenetic mechanisms of mortality risk among PLWH.

We demonstrated that a machine learning approach can predict mortality risk among HIV-positive individuals across different methylation data sets. One of the challenges of a machine learning method is overfitting. We attempted to limit overfitting in the development of the ensemble prediction models in several ways: 1) model development and final model and feature selection were conducted separately in the training and validation sets, 2) 10-fold cross-validation was performed during the training, 3) the ensemble model was used to aggregate prediction results from multiple base models instead of arbitrarily choosing a specific machine learning prediction model, and 4) the final model was evaluated in an independent test set. We further observed that the performance of different models generated by the individual machine learning method varied. The ensemble-based modeling method outperformed some base models and could aggregate prediction results from all base models. Furthermore, our prediction model was built on training and validation sets profiled on the 450K array, and it was independently evaluated in the testing set that was profiled on the EPIC array. Of note, removing batch effects is an important step in EWA analysis. Methods have been applied to address the bias related to batches (62). Here, our goal was to select a set of generalizable methylation markers that are relatively stable regardless of batch, cohort, and other confounding factors. Without removing batch effects within the set of CpG sites common to both arrays, our model still showed good predictive performance, indicating that our model is generalizable, regardless of methylation platform. Our results suggested that the ensemble prediction model is relatively stable and robust.

Our results showed that the CpG-based ensemble prediction model is strongly associated with mortality in an HIV-positive population. This finding is consistent with previous literature showing that DNAm marks in blood can predict mortality in non-HIV populations (40). We found that 30 out of 393 CpGs reached epigenome-wide significance. The majority of the 393 CpG sites are located within or near genes that are involved in known HIV pathology and progression. For example, cg22930808 on *PARP9 and* cg07107453 on *IFI44* were selected by the machine learning prediction model and reached epigenome-wide significance; both genes are involved in HIV pathogenesis. Both cg22930808 and cg07107453 are less methylated in the high mortality risk group than in the low mortality risk group. *IFI44* is an interferon-alfa inducible protein and is associated with infection by several viruses. A previous study showed that higher expression of *IFI44* facilitated HIV-1 latency (63), which may increase mortality risk. In addition, cg12359279 is located in the *MX1* gene (Interferon-Induced GTP-Binding Protein Mx1). *MX1* encodes a GTP-metabolizing protein that is induced by interferon I and II and is involved in interferon gamma signaling and the Toll-like signaling pathway.

Studies have shown that comorbidity and aging are associated with HIV-related excess mortality (64-66), and some immune biomarkers can partially explain HIV-related excess mortality risk (67). Since the VACS index included measures of immune function and indicators for general organ injury, it is plausible that predictive CpG sites (based on the VACS index) are located near genes involved in immune system development or other functions, such as liver or kidney functions. Among the 393 predictive CpG sites, no CpG site located on the genes overlapped with previously reported liver or kidney diseases. However, two of the 393 CpG sites, cg16249932 and cg00463367, are located near genes related to immune system development *(MAEA* and *GATA3)*.

The biological relevance of these 393 CpG sites was further supported by the Gene Ontology enrichment analysis. The top enriched pathways, such as response to virus and cytokine receptor binding, may indicate important biological pathways that lead to increased risk of mortality among PLWH.

We acknowledge that there are several limitations in this study. We defined high mortality risk by a cutoff of 40 for the VACS index based on previous literature, with a predicted 3-year mortality of 10% for individuals with a VACS index score >40 (48); the cutoff for defining high mortality risk may vary in other populations. Additionally, the generalizability of our prediction model may be limited because our samples were predominantly middle-aged men. All samples in our study were HIV-positive, and we cannot infer whether the identified CpGs are relevant to predicting mortality in an HIV-negative population. Last, we are unable to validate the array-based DNA methylation of the selected CpG sites by machine learning or to identify significant CpG sites by EWAS through a different platform. However, in a different sample, we found a high correlation of PBMC methylome between EPIC and the methyl-capture sequencing platform (r=0.986) on the same 4 peripheral blood monocyte cells. Future studies in other populations are warranted to validate our ensemble prediction model and the selected methylation features.

## Conclusions

We identified a panel of 393 predictive DNAm features in blood that was predictive of mortality risk among PLWH. These DNAm features may serve as biomarkers to identify individuals at high risk for mortality and may help to prioritize genes to better understand the mechanisms of mortality risk in an HIV-positive population. These DNAm features have the potential to be used for monitoring HIV progression in future clinical care.

## Data Availability

Data has been deposited in GEO

---

**Figure S1:** Distribution of Veteran Aging Cohort Study (VACS) index between sample sets 1 and 2.

**Figure S2:** Epigenome-wide association (EWA) on mortality risk among PLWH: quantile-quantile plot, Manhattan plot and overlapping CpG sites between EWA significant CpGs and predictive CpGs.

**Table S1:** Predictive CpG sites by variance importance

**Table S2:** Epigenome-wide significant CpG sites in meta-analysis of EWAS on mortality risk among people living with HIV

**List of abbreviations**
AUC: Area Under Curve
CI: Confidence interval
DMR: differentially methylated region
DNA: Deoxyribonucleic acid
DNAm: DNA methylation
DAVID: Database for Annotation, Visualization and Integrated Discovery
EWA: epigenome-wide association
FDR: False discovery rate
FWER: Family-wise error rate
GLMNET: elastic-net regularized generalized linear models
GO: Gene ontology
HIV: Human immunodeficiency virus
HM450K: Human Methylation 450K BeadChip
k-NN: k-nearest neighbors
NK: Natural killer
PC: Principal component
PLWH: people living with HIV
QC: Quality control
SVM: Support Vector Machines
VACS: Veterans Aging Cohort Study
XGBoost: Extreme Gradient Boosting Tree

## Declarations

### Ethics approval and consent to participate

The study was approved by the Committee of the Human Research Subject Protection at Yale University and the Institutional Research Board Committee of the Connecticut Veteran Healthcare System. All subjects provided written consent.

### Availability of data and materials

Demographic and clinical variables and DNAm data for the VACS samples were submitted to GEO dataset (GSE117861) and are available to the public. All codes for analysis are also available upon request to the corresponding author.

### Competing interests

The authors declare that they have no competing interests.

### Funding

This project was supported by the National Institute on Drug Abuse (R03DA039745, R01DA038632, R01DA047063, R01DA047820).

### Authors’ contributions

CS was responsible for data analysis and manuscript preparation. ACJ provided DNA samples and clinical data and contributed to manuscript preparation. XZ was responsible for the bioinformatics data processing. VM was involved in clinical data collection and manuscript preparation. DH and EJ contributed to the analytical approach and to manuscript preparation. KX was responsible for the study design, study protocol, sample preparation, data analysis, interpretation of findings, and manuscript preparation.

## Acknowledgments

The authors appreciate the support of the Veteran Aging Study Cohort Biomarker Core and the Yale Center of Genomic Analysis.

## Notes

### Competing Interest Statement

The authors have declared no competing interest.

### Author Declarations

All relevant ethical guidelines have been followed and any necessary IRB and/or ethics committee approvals have been obtained.

Any clinical trials involved have been registered with an ICMJE-approved registry such as ClinicalTrials.gov and the trial ID is included in the manuscript.

### Summary of Updates

Manuscript has been revised according to reviewer's comments

